# Communicable and non-communicable co-morbidities and the presentation of COVID-19 in an African setting of high HIV-1 and tuberculosis prevalence

**DOI:** 10.1101/2021.05.11.21256479

**Authors:** Elsa du Bruyn, Cari Stek, Remi Daroowala, Qonita Said-Hartley, Marvin Hsiao, Rene T. Goliath, Fatima Abrahams, Amanda Jackson, Sean Wasserman, Brian W Allwood, Angharad G. Davis, Rachel P-J. Lai, Anna K. Coussens, Katalin A. Wilkinson, Jantina de Vries, Nicki Tiffin, Maddalena Cerrone, Ntobeko A. B. Ntusi, Catherine Riou, Robert J. Wilkinson, on behalf of the HIATUS investigators, Saalikha Aziz, Nonzwakazi Bangani, John Black, Marise Bremer, Wendy Burgers, Zandile Ciko, Hanif Esmail, Siamon Gordon, Yolande X. R. Harley, Francisco Lakay, Fernando-Oneissi Martinez-Estrada, Graeme Meintjes, Marc Mendelson, Tari Papavarnavas, Alize Proust, Sheena Ruzive, Georgia Schafer, Keboile Serole, Claire Whitaker, Kennedy Zvinairo

## Abstract

**Objectives:** To describe the presentation and outcome of SARS-CoV2 infection in an African setting of high non-communicable co-morbidity and also HIV-1 and tuberculosis prevalence.

**Design:** Case control analysis with cases stratified by HIV-1 and tuberculosis status.

**Setting:** A single-centre observational case-control study of adults admitted to a South African hospital with proven SARS-CoV-2 infection or alternative diagnosis.

**Participants:** 104 adults with RT-PCR-proven SARS-CoV2 infection of which 55 (52.9%) were male and 31 (29.8%) HIV-1 co-infected. 40 adults (35.7% male, 30.9% HIV-1 co-infected) admitted during the same period with no RT-PCR or serological evidence of SARS-CoV2 infection and assigned alternative diagnoses. Additional *in vitro* data from prior studies of 72 healthy controls and 118 HIV-1 uninfected and infected persons participants enrolled to a prior study with either immune evidence of tuberculosis sensitization but no symptoms or microbiologically confirmed pulmonary tuberculosis.

**Results:** Two or more co-morbidities were present in 57.7% of 104 RT-PCR proven COVID-19 presentations, the commonest being hypertension (48%), type 2 diabetes mellitus (39%), obesity (31%) but also HIV-1 (30%) and active tuberculosis (14%). Amongst patients dually infected by tuberculosis and SARS-CoV-2, clinical features could be dominated by either SARS-CoV-2 or tuberculosis: lymphopenia was exacerbated, and some markers of inflammation (D-dimer and ferritin) elevated in singly SARS-CoV-2 infected patients were even further elevated (p < 0.05). HIV-1 and SARS-CoV2 co-infection resulted in lower absolute number and proportion of CD4 lymphocytes, with those in the lowest peripheral CD4 percentage strata exhibiting absent or lower antibody responses against SARS-CoV2. Death occurred in 30/104 (29%) of all COVID-19 patients and in 6/15 (40%) of patients with coincident SARS-CoV-2 and tuberculosis.

**Conclusions:** In this South African setting, HIV-1 and tuberculosis are common co-morbidities in patients presenting with COVID-19. In environments in which tuberculosis is common, SARS-CoV-2 and tuberculosis may co-exist with clinical presentation being typical of either disease. Clinical suspicion of exacerbation of co-existent tuberculosis accompanying SARS-CoV-2 should be high.

**What is already known on this topic?:** It has been quite widely thought that Africa has been spared the worst effects of the COVID-19 pandemic. There are very few reported case series and no case-control studies comparing COVID-19 patients admitted to hospital to those admitted for other reasons. However several studies have indicated both HIV-1 and tuberculosis co-infection that are endemic in Africa constitute risk factors for poor outcome. In addition Africa is subject to demographic transition and the prevalence of non-communicable co-morbidities such as type 2 diabetes, hypertension and cardiovascular disease is rising rapidly. No study from Africa has described the clinical impact on the presentation of COVID-19 infection.

**What this study adds:** Two or more co-morbidities were present in over half COVID-19 presentations, including HIV-1 (30%) and active tuberculosis (14%). Patients dually infected by tuberculosis and SARS-CoV-2, presented as either SARS-CoV-2 or tuberculosis. HIV-1 and SARS-CoV2 co-infection resulted in lower absolute number and proportion of CD4 lymphocytes, and those with low CD4 counts had absent or lower antibody responses against SARS-CoV2. Death occurred 29% of all COVID-19 patients and in 40% of patients with coincident SARS-CoV-2 and tuberculosis. Thus in environments in which tuberculosis is common, SARS-CoV-2 and tuberculosis may co-exist with clinical presentation being typical of either disease and clinical suspicion of exacerbation of co-existent tuberculosis accompanying SARS-CoV-2 should be high.

## Introduction

In 2020, COVID-19 caused by SARS-CoV-2 infection replaced HIV-1 infection and tuberculosis as the leading global infectious cause of death: 3.19 million by 1st May 2021 ^1^. It is Africa that routinely witnesses the bulk of HIV-1 tuberculosis associated mortality, yet with respect to SARS-CoV-2 the continent has been described as a puzzle ^2^. Fewer cases and deaths from COVID-19 than predicted have been notified. This has been attributed to limited molecular testing (serological data suggests infection has been more widespread ^3 4^), a much younger population (and thus fewer severe cases and deaths), and the possible influences of pre-existing immunity, genetic factors, and the early implementation of public health measures, especially restriction on already limited intracontinental travel.

In the Republic of South Africa, early importation of SARS-CoV-2 into its Western Cape province in March 2020 led, notwithstanding initial very stringent lockdown measures, to a substantial first wave peaking in July 2020 that exerted extraordinary demand on hospital services ^5 6^. Over 1.5 million confirmed infections have occurred, and the South African Medical Research Council estimates 155,859 excess deaths nationally since 3rd May 2020 (https://www.samrc.ac.za/reports/report-weekly-deaths-south-africa). Multiple mutations giving rise to a novel variant referred to as 501Y.V2 contributed to a worse second wave in this region ^7^. The Western Cape province of South Africa has high HIV-1 prevalence and tuberculosis incidence to which considerable and often co-incident non-communicable disease burden is added with obesity, type 2 diabetes mellitus, hypertension, and vascular co-morbidities being very common ^8 9^. Robust provincial health information systems triangulate multiple data sources to enumerate health conditions, with assignment of certainty levels for each enumeration ^10^. These systems were deployed rapidly during the COVID-19 first wave to powerfully identify co-morbidities associated with COVID-19 amongst (i) public sector patients, (ii) laboratory-diagnosed COVID-19 cases and (iii) hospitalized COVID-19 cases. Among 3,460,932 patients (16% HIV-1 infected), 22,308 were diagnosed with COVID-19, of whom 625 (28%) died. In addition to widely recognised risk factors (male sex, increasing age, diabetes, hypertension and chronic kidney disease) COVID-19 death was associated with HIV-1 (adjusted hazard ratio [aHR] 2.14), with similar risks irrespective of viral load and immunosuppression, and also current and previous tuberculosis (aHR 2.70 and 1.51 respectively ^11^).

A number of clinical studies have examined interaction between HIV-1 and SARS-CoV-2: mostly cohorts lacking HIV-1- or SARS-CoV-2-uninfected control participants ^12-21^. Definitive conclusions are thus difficult to reach but the presentation and outcome of SARS-CoV-2 in HIV-1 co-infected patients is reported as not differing greatly from that in HIV-1 uninfected persons ^22-24^. A single report associated the use of tenofovir as disoproxil fumarate with better outcome than as alafenamide ^18^. Two reports suggest worse outcomes associated with lower CD4 count ^17 25^ and a further report suggests prolonged SARS-CoV-2 shedding (determined by RT-PCR) and reduced anti-SARS-CoV-2 antibody response in those whose antiretroviral therapy (ART) has been interrupted ^26^. Interestingly convalescence from COVID-19 appears associated with a transient increase in HIV-1 viral load despite ART ^19 27^.

For tuberculosis, even less systematic information exists. Motta *et al*. reported coincident tuberculosis in their own, and a prior, cohort of, co-infections concluding tuberculosis might not be a major determinant of mortality ^28 29^. Stochino *et al*. reported a case series of what appears to be nosocomial transmission of SARS-CoV-2 amongst tuberculosis inpatients noting tuberculosis outcomes to be generally unaffected ^30^. Interestingly in a study of 159 South African children with SARS-CoV-2, 51 of 62 hospitalised were symptomatic and of these 7/51 had a recent or current diagnosis of tuberculosis ^31^. In a meta-analysis heavily influenced by the aforementioned population-based study by Boulle *et al*. ^11^, Sarkar and colleagues concluded patients with tuberculosis have an increased risk of mortality during co-infection with SARS-CoV-2 (risk ratio = 2.10; 95% CI, 1.75-2.51) ^32^. More recently a postmortem surveillance study conducted in Zambia reported the presence of tuberculosis in 22/71 (31%) patients who died with a positive RT-PCR for SARS-CoV-2 ^33^.

We therefore instigated a series of health facility-based observational studies to investigate interaction and overlap between SARS-CoV-2, HIV-1 and *M. tuberculosis* infections (HIATUS). The purpose was to describe the clinical presentation, radiographic appearances, and clinical laboratory features of patients hospitalized with proven or suspected SARS-CoV-2 infection in an African setting not only endemic for HIV-1 and tuberculosis but also subject to demographic transition such that other non-communicable co-morbidities are common ^9^.

## Methods

### Participant enrolment

This was a single-centre observational case-control study of adult (age > 18 years) patients admitted to Groote Schuur Hospital, Cape Town, South Africa with RT-PCR proven, or suspected, SARS-CoV-2 infection. Between 11^th^ June and 28^th^ August 2020, the study team assessed suspected or confirmed SARS-CoV-2 (by nasopharyngeal or oropharyngeal aspirate) participants and expedited written electronic consent for inclusion was provided. Where participants did not have capacity to provide informed consent, e.g. in the case of those admitted in the intensive care unit, informed consent was sought from family members. Informed consent was provided by all participants who regained their capability to provide informed consent after incapacity. Known HIV-1 status or willingness to undergo testing was an inclusion criterion. During the initial surge of hospital admissions recruitment could not be sequential due to caseload. Emphasis was on known SARS-CoV-2 PCR positive inpatients in a variety of dependency settings ranging through person under investigation (PUI), low dependency known positive, high-flow nasal oxygen and intensive care settings. As caseload decreased emphasis on PUI recruitment increased such that appropriate inpatient controls without laboratory evidence of SARS-CoV-2 infection (other disease controls, ODC) could be recruited. Data were entered directly into an electronic REDCap data entry system, hosted by the University of Cape Town ^34^. The initial aim was to recruit equal numbers of HIV-1-co-infected and -uninfected participants. Participants presenting as persons under investigation (PUI) for SARS-CoV-2 but persistently negative by RT-PCR (Seegene, Roche or Gene Xpert) for SARS-CoV-2 were subsequently serologically tested. In the absence of either RT-PCR or serological evidence of SARS-CoV-2 infection and in the presence of an alternative diagnosis such participants were designated controls. The WHO ordinal severity score was used to categorize participants on admission according to the following categories:

1. Not hospitalized, no limitation on activities;
2. Not hospitalized, limitation on activities;
3. Hospitalized, not requiring supplemental oxygen;
4. Hospitalized, requiring supplemental oxygen by mask or nasal cannulae
5. Hospitalized, on non-invasive ventilation or high flow oxygen devices;
6. Hospitalized, on invasive mechanical ventilation;
7. Hospitalized, on invasive mechanical ventilation with other organ support.
8. Death

The study was conducted according to the declaration of Helsinki, conformed to South African Good Clinical Practice guidelines, and was approved by the University of Cape Town’s Health Sciences Research Ethical Committee (HREC 207/2020).

For some analyses we included data from HIV-1 uninfected and infected persons with either immune evidence of tuberculosis sensitization but no symptoms (latent tuberculosis, LTBI) or microbiologically confirmed pulmonary tuberculosis who had been recruited to prior studies (HREC 050/2015) ^35 36^. The details of these persons are presented in Supplemental Table 2.

### Laboratory evaluations

Serological testing was performed by Elecsys® (Roche, Basel, Switzerland) Anti-SARS-CoV-2 immunoassay which detects antibodies (including IgG) to SARS-CoV-2. The following routine clinical assays were performed in the laboratories of the National Health Laboratory Service (NHLS): full blood count and automated differential cell count, HIV-1 ELISA, Microbiology including tuberculosis Gene Xpert nucleic acid amplification testing, HIV-1 viral load, peripheral blood CD4 percentage and absolute count, SARS-CoV-2 diagnostic PCR (Seegene, Roche, Gene Xpert), blood electrolytes, C-reactive protein, D-dimer, ferritin and lactate dehydrogenase. The NHLS via its quality assurance division provides proficiency testing to its own and other laboratories schemes and is certified to ISO/IEC 17043:2010 requirements.

### Flow cytometric analysis

The assessment of the percentage of CD4 and CD8 T cells was performed on cryopreserved fixed whole blood. Briefly, samples were thawed, washed and permeabilized with a Transcription Factor perm/wash buffer (eBioscience, San Diego, CA, USA). Cells were then stained with an antibody cocktail containing among others anti-CD3, -CD4 and -CD8 antibodies. Samples were acquired on a BD LSR-II and analyzed using FlowJo (v9.9.6, FlowJo LCC, Ashland, OR, USA). The percentage of CD4 and CD8 T cells were defined as a % of total lymphocytes identified based on their FSC and SSC characteristics.

### Radiographic evaluation

Chest radiographs were reported blind to clinical status by an experienced radiologist (QS-H). The BSTI coding system was employed to report chest radiographs as described ^37^ and the Brixia severity score was also calculated as described ^38^. Posteroanterior chest radiographs were assessed for the total percentage of the lung fields unaffected by any visible pathology. Thus, in the COVID-19 group this score quantified the percentage of normal lung that was not visibly affected by known features of COVID-19 pneumonia on the radiograph. In those with tuberculosis, or in ODC with other respiratory infections this score similarly quantified the percentage of normal lung, not visibly affected by the relevant pathology on the radiograph. Those with a normal chest radiograph would thus score 100%.

### Analytical methods

Analyses were conducted in Prism 8 (GraphPad Software, San Diego, CA). The normality of data was assessed by a Shapiro-Wilk test. Descriptive statistics were calculated and presented as percentage or median and IQR. Unpaired normally distributed variables were compared by the student’s unpaired t test and non-parametric comparisons by the Kruskal-Wallis test. Corresponding assessment of non-parametric correlation was by the Spearman method.

### Patient and public involvement

- At what stage in the research process were patients/public first involved in the research and how? It was difficult to engage relevant communities in the design of the research as it was conceived during a period of stringent national lockdown during which unnecessary gatherings and travel were illegal.
- How were the research question(s) and outcome measures developed and informed by their priorities, experience, and preferences? In a country in which around 1 in 6 adults are HIV-1 infected there was immense public interest in how pre-existing HIV-1 infection might modify the outcome of SARS-CoV2 infection
- How were patients/public involved in the design of this study? As above
- How were they involved in the recruitment to and conduct of the study? Recruitment was shortly after admission to hospital with suspected SARS-CoV2 infection and accompanied by an electronic informed consent procedure with proxy consent (and subsequent reconsent for those incapacitated). For those who did not regain capacity or died the research ethical committee was consulted to determine if such data could be included on a case-by-case basis.
- Were they asked to assess the burden of the intervention and time required to participate in the research? Yes, a consent procedure as outlined above was undertaken. The research was designed to minimally burden participant and the service during the time of an emergency.
- How were (or will) patients and the public be involved in choosing the methods and agreeing plans for dissemination of the study results to participants and wider relevant communities? Results have already been shared via academic and lay fora. The lesson that HIV-1 and tuberculosis may co-exist in SARS-CoV2 patients has been widely disseminated and appreciated.

## Results

104 SARS-CoV-2 RT-PCR positive and 50 negative participants (non-COVID-19 other disease controls, ODC) were enrolled. Eight of the 50 RT-PCR negative ODC participants tested SARS-CoV-2 antibody positive and were thus excluded from further analysis. The baseline characteristics of the remaining 146 participants are shown in Table 1, with the final diagnosis of ODC participants being shown in Supplemental Table 1. COVID-19 cases and ODC were similar in age, sex, percentage HIV-1 or tuberculosis co-infected (29.8% and 14.4% versus 30.9% and 11.9% respectively) and days of illness prior to blood sampling. HIV-1 co-infected ODC had a lower CD4 count (median: 18 cells/mm^3^ [IQR: 7-102] versus 132 [51-315], p=0.028) and higher median HIV-1 viral load (median: 5.36 log10 HIV RNA copies/ml [IQR: 2.49-5.21] versus <1.3 [<1.3-4.14], p=0.0005). Although the overall frequency of co-morbidities was similar between COVID-19 patients and ODC (Supplemental Figure 1), ODC were more likely to exhibit cardiovascular or other respiratory comorbidities (6.7% and 6.7% versus 42.9 and 26.2%, respectively). The majority of participants in both groups had two or more co-morbidities (COVID-19 57.7% and ODC 74.4%). 46.2% COVID-19 patients were assessed as grade 5 or above on the WHO ordinal scale for COVID-19. There was greater use of adjunctive corticosteroid therapy (78.8% versus 26.1%, p < 0.0001) and a longer hospital stay (13 versus 7 days, p=0.0008) in COVID-19 patients.

**Table 1.** Clinical characteristics of COVID-19 versus hospitalized controls.

|  | COVID-19 (n=104) | non-COVID-19 ODC (n=42) | p-value |
| --- | --- | --- | --- |
| Age (median, IQR) | 53 [44-61] | 51 [36-66] | 0.55 |
| Male (%) | 52.9% (n=55) | 35.7% (n=15) | 0.06 |
| HIV-1 co-infected (% , n) | 29.8% (n=31) | 30.9% (n=13) | 0.89 |
| On antiretroviral therapy (ART) | 74.2% (n=23) | 46.1% (n=6) | 0.073 |
| Time on ART (years) <sup>a</sup> | 9.6 [6-12] | 3.6 [0.4-11] | 0.11 |
| CD4 count (cells/mm<sup>3</sup>) <sup>a</sup> | 132 [51-315] | 18 [7-102] | 0.028 |
| CD4 count (cells/mm<sup>3</sup>) in ART established | 188.5 [76-336] | 137 [5-780] | 0.804 |
| CD4 count (cells/mm<sup>3</sup>) in ART naïve | 66 [14-106] | 18 [7-55] | 0.24 |
| Log<sub>10</sub> HIV Viral load <sup>a</sup> | <1.3 [<1.3-4.14] | 5.36 [2.49-5.52] | 0.0005 |
| Log<sub>10</sub> HIV viral load in ART established | <1.3 [<1.3-<1.3] | 3.08 [<1.3-5.4] | 0.098 |
| Log<sub>10</sub> HIV viral load in ART naïve | 4.44 [4.13-4.8] | 5.44 [5.0-5.56] | 0.004 |
| *M. tuberculosis* positive (% , n) | 14.4 % (n=15) | 11.9 % (n=5) | 0.69 |
| Previous tuberculosis episode/s (within 5 years) | 6.7% (n=7) | 9.5% (n=4) | 0.73 |
| Co-morbidities (% , n) |  |  |  |
| Cardiovascular | 6.7% (n=7) | 42.9% (n=18) | <0.0001 |
| Hypertension | 48.1% (n=50) | 54.8% (n=23) | 0.46 |
| Diabetes | 39.4% (n=41) | 28.6% (n=12) | 0.22 |
| Obesity | 32.7% (n=34) | 33.3% (n=14) | 0.98 |
| Other respiratory diseases | 6.7% (n=7) | 26.2% (n=11) | 0.001 |
| SARS-CoV-2 PCR positive | 100% | 0% | - |
| SARS-CoV-2 serology positive <sup>b</sup> | 69.2% (n=72) | 0% | - |
| Cut-off index (median, IQR) <sup>c</sup> | 7.06 [0.38-25.78] | 0.07 [0.07-0.08] | <0.0001 |
| WHO COVID-19 ordinal scale at enrolment (% , n) |  |  |  |
| 2 | - | 16.7% (n=7) | - |
| 3 | 17.3% (n=18) | 42.8% (n=18) | 0.001 |
| 4 | 36.5% (n=38) | 40.5% (n=17) | 0.66 |
| 5 | 28.8% (n=30) | - | - |
| 6 | 16.3% (n=17) | - | - |
| 7 | 0.96% (n=1) | - | - |
| Mild and moderate (WHO <5, %) | 53.8% (n=56) | 100% | <0.0001 |
| Severe (WHO ≥5, %) | 46.2% (n=48) | 0% | - |
| On steroid treatment (% , n) | 78.8% (n=82) | 26.1% (n=11) | <0.0001 |
| Thrombo-embolic complications (% , n) | 9.6% (n=10) | 0% | - |
| Days with symptoms at sampling <sup>a</sup> | 9 [6-14] | 8 [3-18] | 0.75 |
| Overall days in clinical care <sup>a</sup> | 13 [7-24] | 7 [4-13] | 0.0008 |

ODC: Other disease controls, IQR: Interquartile range,
<sup>a</sup> Median and [IQR] <sup>b</sup> Two values were missing <sup>c</sup> SARS-CoV-2 serology was performed using the Roche Elecsys assay, measuring SARS-CoV-2 nucleocapsid-specific antibodies. Results are reported as numeric values in form of a cut-off index (signal sample/cut-off), where a COI < 1.0 corresponds to non-reactive plasma and COI ≥ 1.0 to reactive plasma.

The analysis was neither designed nor powered to determine risk factors for death amongst COVID-19 patients. 30 deaths occurred (29% of the total diagnosed SARS-CoV-2 RT-PCR positive) of which 23 (77%) were males, the median age was 55 years (IQR, 46-66), 25 (83%) were classified as WHO grade 5 or above, and 28 (93%) received corticosteroid therapy (Supplemental Table 2). 20% patients who died were HIV-1 co-infected, 20% had tuberculosis and 10% had the triple combination of SARS-CoV-2, tuberculosis and HIV-1. 6/15 (40%) of patients with coincident tuberculosis died of whom equal numbers were HIV-1 infected and uninfected (Supplemental Tables 2 and 3).

Chest radiographs at enrolment were evaluated by three scoring systems: Brixia ^38^, British Society for Thoracic Imaging (BSTI) ^37^ and the percentage of unaffected lung. There was a close relationship between Brixia and BSTI radiographic severity scores (Supplemental Figure 2A); and an inverse relationship between both the BSTI and Brixia scores and the percentage of unaffected lung (supplemental figures 2B and C). The percentage of unaffected lung was lower in COVID-19 patients than ODC (Median 35 vs 70%, p < 0.0001, Supplemental Figure 2D). There was no significant effect of either HIV-1 or tuberculosis co-infection alone on the percentage unaffected lung in COVID-19 patients, but patients with triple infection tended to exhibit a higher percentage of unaffected lung (p = 0.007, Supplemental Figure 2E).

A complete listing of patients with both tuberculosis and COVID-19 (+/- HIV-1 infection) is provided in Supplemental Table 3 and examples of radiographic appearances are provided in Figure 1A and B and Supplemental Figure 3A-D. In COVID-19 patients, both Brixia and BSTI radiographic scores related closely to the WHO clinical severity score (Figure 1D and E). There was a trend towards a greater proportion of radiographs classified as non-COVID-19 like in those with COVID-19 and coincident tuberculosis, regardless of HIV-1 status (Figure 1C). In HIV-1-co-infected, tuberculosis-positive, SARS-CoV-2 positive patients the Brixia score was lower than both singly SARS-CoV-2 positive patients and HIV-1 uninfected, tuberculosis-positive, SARS-CoV-2 positive patients (median score: 8 vs. 14 and 15.5, p = 0.006 and 0.038, respectively, Figure 1F). These data therefore indicate that in persons SARS-CoV-2 RT-PCR positive, radiographic appearances were most often typical of COVID-19 with the finding of tuberculosis being incidental, but that in a smaller number of cases appearances were predominantly of tuberculosis, with SARS-CoV-2 being co-prevalent. HIV-1 is known to be associated with non-specific radiographic appearances of tuberculosis ^39^ and appears in triple combination to associate with atypical appearances also of COVID-19 that were ostensibly less severe.

**Figure 1:**
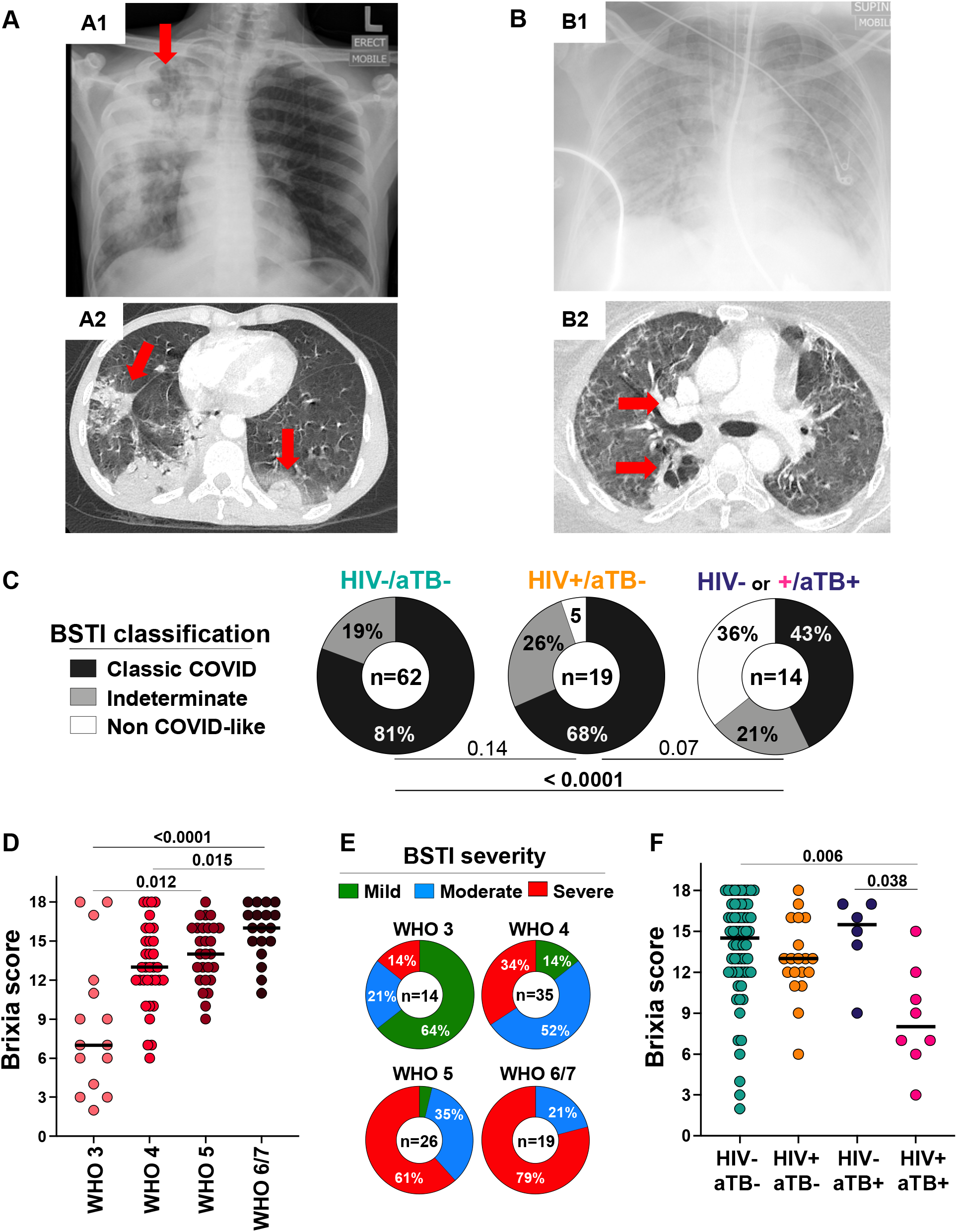
Radiographic appearances of combined SARS-CoV-2 and *M. tuberculosis* infection and relationship between radiographic (Brixia and British Society for Thoracic Imaging (BSTI)) score and clinical severity assessed by WHO COVID-19 ordinal scale. **A&B** Chest radiographs and computed tomographic pulmonary angiography (CTPA) of two COVID-19 patients. **A1:** 40-45-year-old HIV-1 uninfected patient presenting with consolidation and cavitation in the right upper lobe (red arrow), 3+ sputum smear positive and also SARS-CoV-2 RT-PCR positive (threshold cycle: 32.45). **A2:** Because of persistent hypoxia and tachypnoea the patient underwent CTPA which showed bi-basal wedge-shaped opacities in keeping with pulmonary embolism (PE) and/or consolidation related to COVID-19. Discharged after 22 days. **B1:** 40-45-year-old HIV-1 uninfected patient presenting with SARS-CoV-2 positive RT-PCR (threshold cycle: 21.2) with diffuse bilateral shadowing on the chest radiograph who deteriorated necessitating intubation and ventilation for 36 days. The patient remained O_2_ dependent after extubation and a CTPA (**B2**) for suspected PE instead revealed cavitation associated with opacification and air bronchograms in the superior segment of the right lower lobe together with subcarinal and pretracheal lymphadenopathy. The patient was found Gene Xpert MTB/Rif positive. **C** BSTI classification of RT-PCR proven SARS-CoV-2 cases in the absence or presence of HIV-1 and/or tuberculosis. The increase in the proportion of radiographs classified as Non-COVID like tended to increase in those with coincident tuberculosis. Comparisons were performed by Chi-square test **D** Brixia radiographic and, **E** BSTI, severity scores in relation to WHO clinical severity scale. Comparisons were performed using a Kruskal-Wallis test. **F** Brixia score in relation to the presence or absence of HIV-1 and/or tuberculosis co-infection. The extent of changes related COVID-19 was decreased amongst those with coincident HIV-1 associated tuberculosis. Comparisons were performed using a Kruskal-Wallis test with Dunn’s correction.

### Peripheral white cell count

Peripheral blood data stratified by COVID-19 status and WHO severity (3-7) are shown in Figure 2. Data from 72 HIV-1 uninfected healthy controls (HC) recruited to pre-pandemic studies (pre-2019) were also used for comparison (Supplemental table 4). The total white cell count was raised in both ODC and COVID-19 patients by comparison with HC with no differences between the patient groups, and the count increased with increasing COVID-19 severity (Figure 2A). Amongst COVID-19 patients, the total white cell count was highest in HIV-1 uninfected patients with coincident tuberculosis (median: 15.34 x 10^9^ /L, IQR: 11.97-17.13). Lymphocyte counts were lower in both ODC and COVID-19 patients (with no differences between these groups) compared to HC. The count trended towards a decrease with increasing COVID-19 severity and was significantly lowest in triply infected patients (p = 0.03, Figure 2B). Both neutrophil and monocyte counts were raised in both ODC and COVID-19 patients with no differences between these groups. Neutrophil, but not monocyte, count related to increasing COVID-19 severity (Figure 2C&D). Those with coincident tuberculosis had higher monocyte counts than those with combined HIV-1 and COVID-19 infections without tuberculosis (p = 0.04, Figure 2D). By comparison with HC, eosinophil counts were lower in both ODC and COVID-19 patients were lowest in those with COVID-19 infection with a trend observed in relation to disease severity (p = 0.004, Figure 2E).

**Figure 2:**
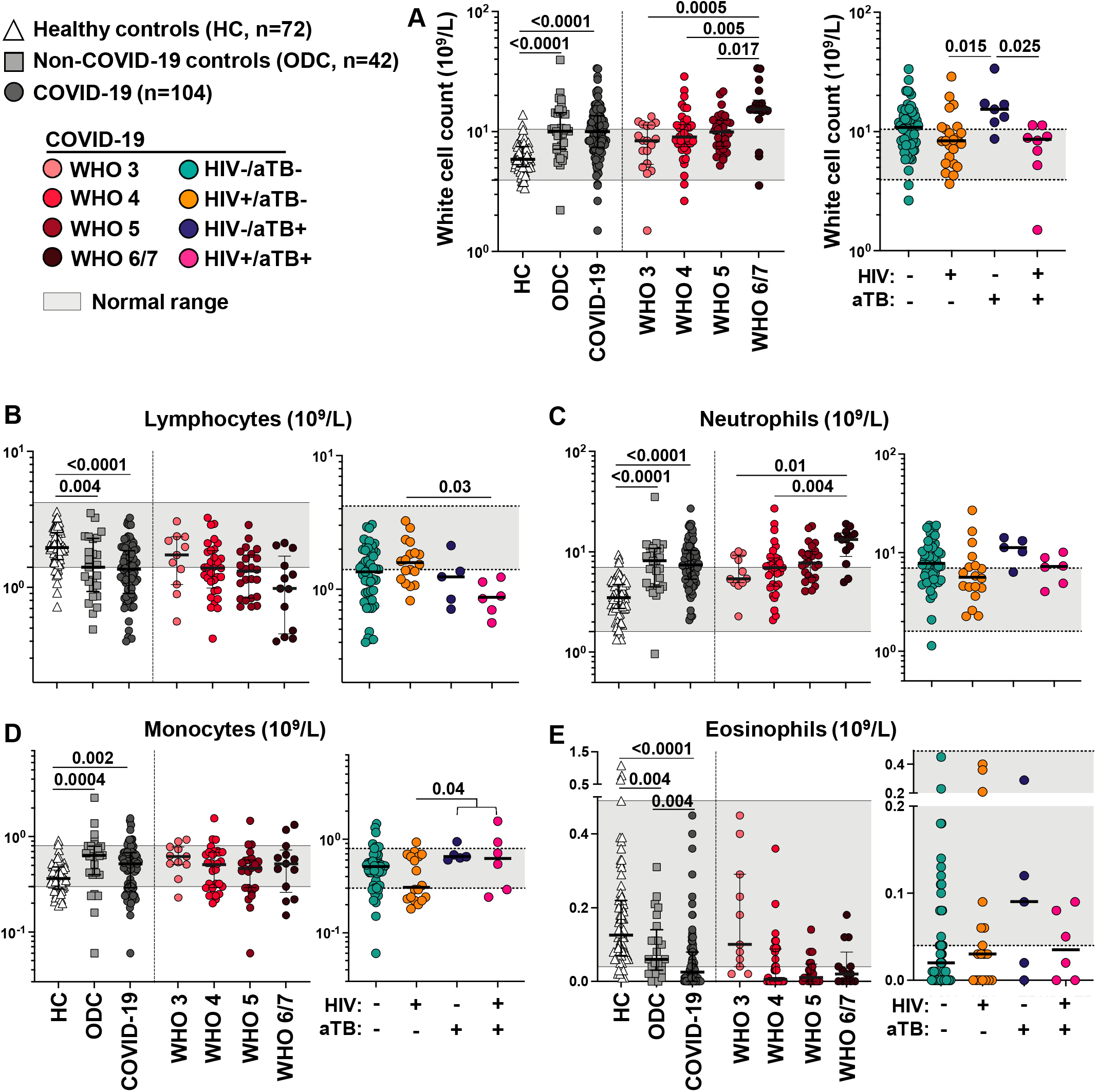
Total white cell and differential count in healthy persons (HC), non-COVID-19 hospitalized controls (ODC), and COVID-19 participants. **A** The total white cell count was raised in both COVID-19 and ODC patients with no differences between the groups. The count increased with increasing COVID-19 severity. Amongst COVID-19 patients the total white cell count was highest in HIV-1 uninfected patients with coincident tuberculosis. **B** The lymphocyte count was lower in both COVID-19 and ODC patients with no differences between these groups. The count tended to decrease with increasing COVID-19 severity. Amongst COVID-19 patients the lymphocyte count was lowest in triply infected patients with HIV-1 and coincident tuberculosis. **C** The neutrophil count was raised in both ODC and COVID-19 patients with no differences between the groups. The count increased with increasing COVID-19 severity. Amongst COVID-19 patients there were no differences in count in the presence of HIV-1 and/or tuberculosis. **D** The monocyte count was raised in both ODC and COVID-19 patients with no differences between these groups. No trend in relation to COVID-19 severity was observed. Amongst COVID-19 patients, those with coincident tuberculosis had higher counts than those with combined HIV-1 and COVID-19 infections without tuberculosis. **E** Eosinophil counts were lower in both COVID-19 and ODC patients being lowest in those with COVID-19 infection with a trend in this decrease observed in relation to disease severity. Amongst COVID-19 patients, no significant effect of HIV-1 and/or tuberculosis infection was observed. All comparisons were performed using a Kruskal-Wallis test with Dunn’s correction.

We further assessed the nature of lymphopenia by flow cytometric analysis of whole blood. There was strong correlation between absolute CD4 numbers and the % CD4 determined by flow cytometry (Supplemental Figure 4A). Amongst COVID-19 patients increasing disease severity correlated with a decrease in CD4 and CD8 positive lymphocytes (Supplemental Figure 4B). To investigate lymphocyte differences in COVID-19 patients, additional values were obtained from a subset of 118/163 similar ambulant participants enrolled to a prior study of HIV-1 uninfected and infected persons with either immune evidence of tuberculosis sensitization but no symptoms or microbiologically confirmed pulmonary tuberculosis treated at a community health clinic (Supplemental Table 4 and ^35 36^). When compared to HIV-1 uninfected healthy persons, the percentage of lymphocytes positive for CD4 was lower in HIV-1 uninfected ODC (p ≤ 0.03) and more so in COVID-19 patients (p < 0.0001, Supplemental Figure 4C). Amongst HIV-1 infected patients the values were especially low for ODC. Amongst patients with coincident HIV-1 and tuberculosis infection, % of CD4 positive lymphocytes was also very low irrespective of the presence or absence of SARS-CoV-2 infection. When compared to HIV-1 uninfected HC, the percentage of lymphocytes positive for CD8 was also lower in both HIV-1 uninfected ODC and more so in COVID-19 patients (Supplemental Figure 4C). However, this pattern was not observed amongst HIV-1 infected patients. The % of CD8 positive lymphocytes was markedly depressed in HIV-1 uninfected patients with coincident tuberculosis and SARS-CoV-2 infection (median: 11.2%, IQR: 10.2-15.1). Amongst HIV-1 uninfected patients with COVID-19 there was a significant trend towards a lower percentage of lymphocytes positive for CD4 and CD8 (Supplemental Figure 4B) with increasing WHO grade severity.

### Serum biomarkers

A number of soluble biomarkers have been associated with COVID-19 and its severity ^40^. In our study, higher C-reactive protein, D-dimer, lactate dehydrogenase (LDH) and ferritin levels were observed in COVID-19 patients compared to ODC (Figure 3A-D). In the case of ferritin, this difference approached 10-fold higher (1116 μg/L (IQR: 643-1974) vs 127 μg/L (IQR: 77-477), respectively, p < 0.0001). However, apart from LDH (Figure 3D), there was no significant trend in any of these markers in relation to COVID-19 severity. HIV-1 infected patients with both COVID-19 and tuberculosis had higher levels of D-dimer and ferritin than those HIV-1 infected with COVID-19 alone. Amongst COVID-19 patients there was strong positive correlation between total white cell (WCC) and neutrophil counts (rho = 0.95, p < 0.0001); moderate correlation between ferritin and LDH, and between lymphocyte and monocyte counts (Figure 3E). The relationship between (WCC) and neutrophil counts, and between ferritin and LDH was also observed in ODC (Figure 3F).

**Figure 3:**
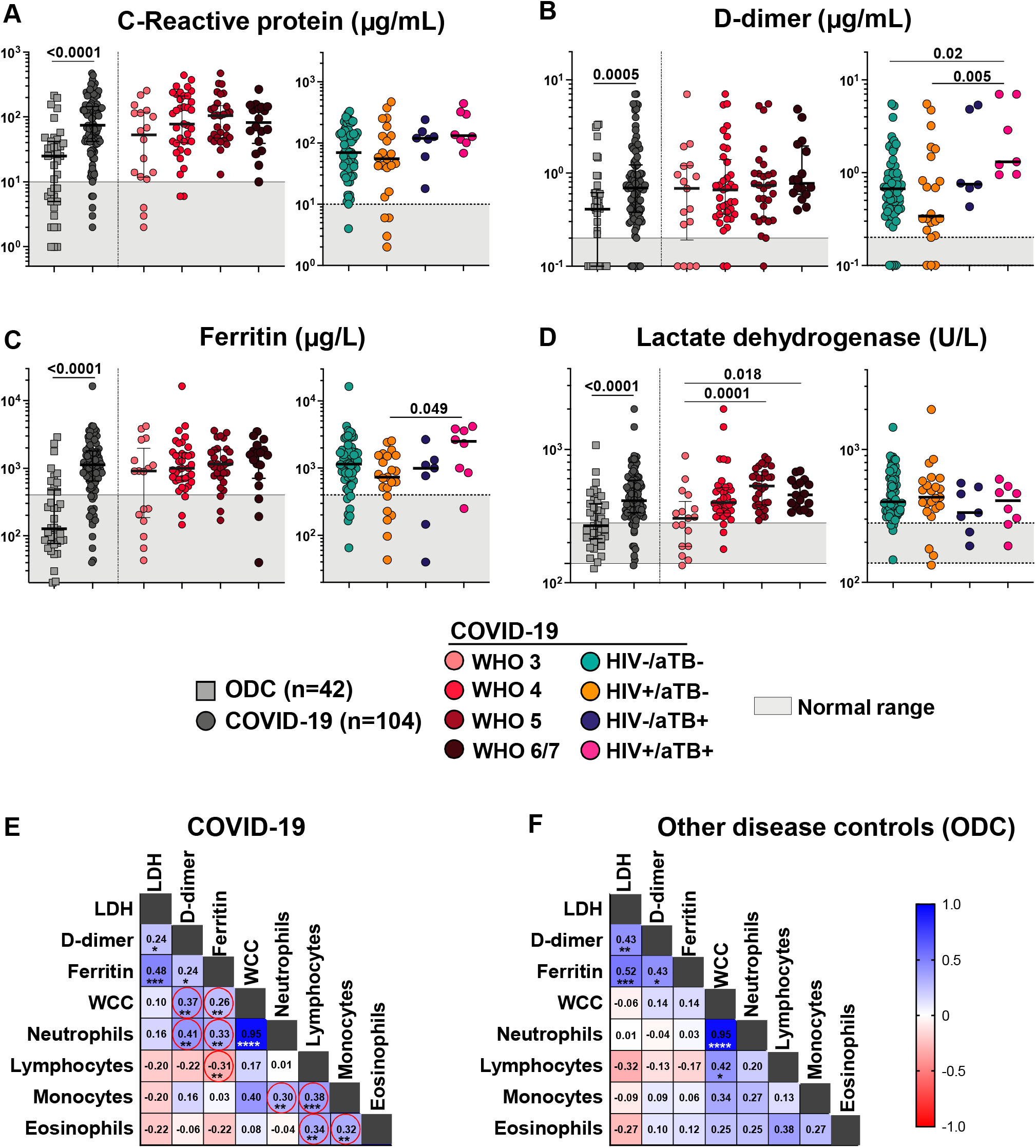
Serum biomarkers in relation to disease status and severity, and correlation between those markers and peripheral cell counts. **A-C** C-Reactive protein (CRP), D-dimer and Ferritin levels showed similar trends being significantly higher in COVID-19 patients than ODC. There was no significant trend in relation to severity of COVID-19 disease. HIV-1 infected patients with both COVID-19 and tuberculosis had higher levels of all three markers than those with COVID-19 alone. **D** Lactate dehydrogenase was significantly higher in COVID-19 patients than ODC, with levels tending to increase with increasing COVID-19 disease severity. There was however no relationship with HIV-1 and/or tuberculosis status. Comparisons between the ODC and COVID-19 groups were performed using a Mann Whitney test and other comparisons were performed using a Kruskal-Wallis test. **E&F** Correlation matrix of serum biomarkers and white cell counts in COVID-19 patients (E) and ODC participants (F). Amongst COVID-19 patients there was strong positive correlation between total white cell (WCC) and neutrophil counts, and moderate between ferritin and LDH, and between lymphocyte and monocyte counts. In ODC participants, the relationship between total white cell (WCC) and neutrophil counts, and between ferritin and LDH was also observed. Positive correlations are indicated in blue and negative correlation in red. Spearman R-values are indicated in each square. \**P* < 0.05, \*\**P* < 0.01, \*\*\**P* < 0.001, \*\*\*\**P* < 0.0001

### SARS-CoV-2 specific antibody response

We next determined relationships between disease severity amongst COVID-19 patients, the presence of HIV-1 and/or tuberculosis as co-morbidities, and the level of anti-SARS-CoV-2 nucleocapsid specific antibody levels determined by the Roche Elecsys assay ^41^. We have previously reported increased antinucleocapsid antibody levels in COVID-19 patients with more severe disease although no overall association with survival was noted ^42^. Although no significant quantitative difference in cut-off index was observed by HIV-1 and or tuberculosis status, 8/15 tuberculosis patients in total and 5/8 with HIV-1 and tuberculosis were below the threshold for positivity (Figure 4A). There was a significant trend towards decreased antibody levels in participants with a lower percentage CD4 lymphocytes with just 3/10 antibody responders amongst patients with a CD4 percentage of lymphocytes below 10% (Figure 4B). Amongst HIV-1 co-infected COVID-19 patients, there was positive correlation between the percentage of CD4 T cells and the antibody cut-off index with triply infected patients tending to show low levels of both parameters (Figure 4C).

**Figure 4:**
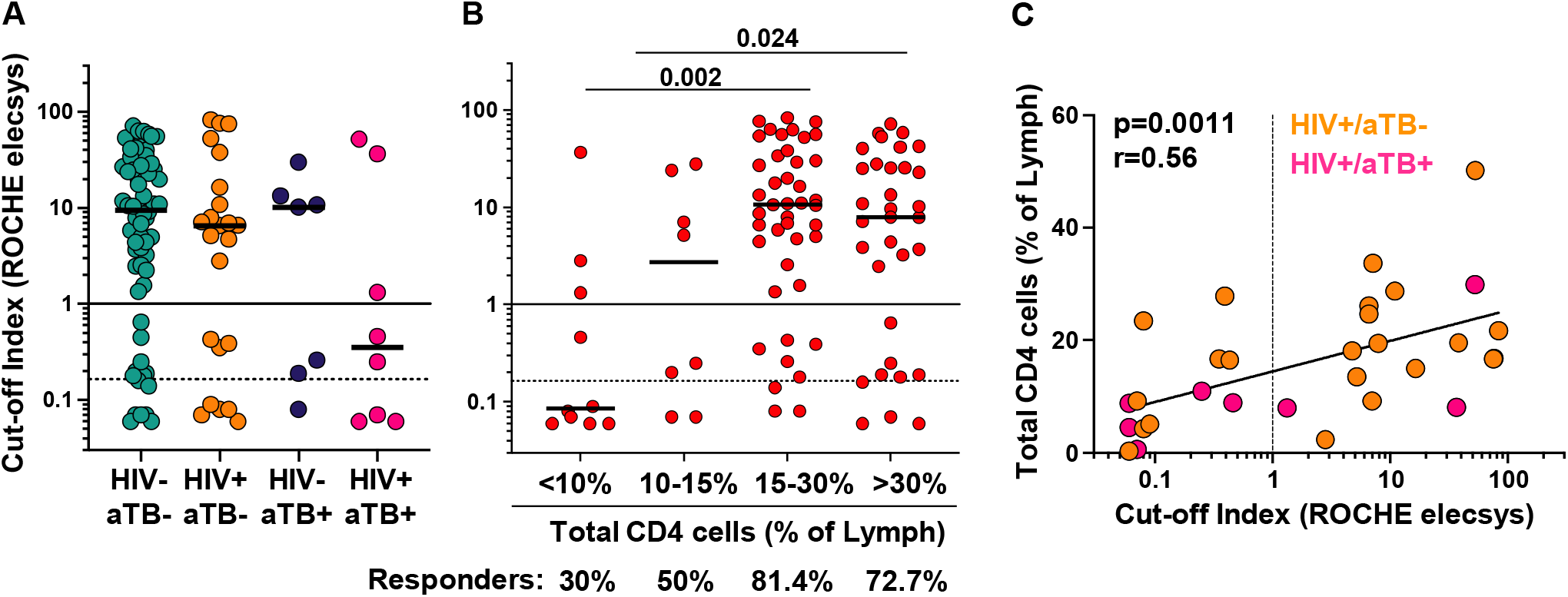
Serum anti-nucleocapsid IgG levels in COVID-19 patients in relation to the presence of HIV-1 and/or tuberculosis co-infection. **A** Although no significant quantitative difference in cut-off index was observed by HIV-1 and or tuberculosis status, 8/15 tuberculosis patients in total and 5/8 with HIV-1 and tuberculosis were below threshold for positivity of 1.0. The >1 cut-off value represents the manufacturer’s cut-off. The >0.165 cut-off value represents the optimal cut-off value defined by ROC curve analyses on 197 participants that improved the performance of the test to give a sensitivity of 100% (95%CI: 94.0-100%) (Reference 45). **B** There was a significant trend towards decreased antibody levels in participants with Lower percentage CD4 lymphocytes. Comparisons were performed using a Kruskal-Wallis test. **C** Amongst HIV-1 co-infected COVID-19 participants, there was positive correlation between the percentage of CD4 lymphocytes and the antibody cut-off index with triply infected patients tending to show low levels of both parameters. Correlation was tested by a two-tailed non-parametric Spearman rank test.

## Discussion

By means of a case-control analysis, we have set out one of the first accounts of the clinical presentation and laboratory features of COVID-19 in an African setting in which there is high non-communicable, but also communicable (HIV-1 and tuberculosis) co-morbidity. An important conclusion is that the triple occurrence of SARS-CoV-2, HIV-1 and tuberculosis is not uncommon. Clinical features may be dominated by either SARS-CoV-2 or tuberculosis and, although overall radiographic severity may be modulated, lymphopenia is exacerbated, and some markers of inflammation are even more elevated in such patients. Forty percent of patients with coincident COVID-19 and tuberculosis died.

Our study benefitted inclusion of participants (ODC) admitted to the same hospital at the same time as the surge in COVID-19 admissions who were subsequently assigned alternative diagnoses with negative virological and serological tests for SARS-CoV-2. We cannot exclude under the circumstances that such tests were false negative. However, the existence of this control group allowed us to compare blood markers that have been used to characterise and prognosticate SARS-CoV-2 infection. We found that neither raised total white count, neutrophilia or lymphopenia can be inferred as specific for COVID-19 (Figure 2). By contrast, the plasma levels of several markers including the CRP, D-dimer, LDH and especially ferritin were markedly elevated in COVID-19 patients (Figure 3). Of these markers however, only LDH showed an increasing trend with COVID-19 disease severity. A potential confounder of results on peripheral blood markers is the greater use of corticosteroid therapy at the time of sampling in COVID-19 patients. The results of the Recovery trial were announced shortly after we began recruitment to our study and were accompanied by an overnight change in clinical guidance to prescribe such therapy to those severely ill ^43^. Notwithstanding this potential blunting of values, our results again reflect the marked severe systemic effects of acute SARS-CoV-2 infection? that are now very widely documented. A greater proportion of participants who died also received steroids. This therapy is indicated in severe COVID-19 ^43^ and this association is therefore probably confounded by the fact that more severely ill participants were more likely to receive this treatment.

Our analysis tends towards the conclusions of others that have included HIV-1 infected persons in not documenting major differences in presentation and outcome except in those profoundly immunosuppressed. ART use was variable and not equally distributed between COVID-19 cases and ODC with the latter more likely to be ART-naïve: perhaps reflecting late presentations of HIV-1 infection at a time when pressure on health services was acute. This feature also precludes meaningful and well-powered comparisons according to ART status. Overall, HIV-1 infected patients recruited to this study were not more likely to die. There was a predictable decrease in CD4 T cell numbers and percentage in HIV-1 infected participants, not reflected however in CD8 counts. Of potentially greater significance, in terms of longer lasting immunity in convalescence, there was a direct relationship between the peripheral CD4 percentage and acute antibody levels. Whilst this may reflect duration of COVID-19 illness and its severity (both tending to increase antibody levels), it remains a grave concern that both natural and vaccine-induced immune responses to SARS-CoV-2 may be less effective in persons immunosuppressed by HIV-1, irrespective of ART status (https://ir.novavax.com/news-releases/news-release-details/novavax-covid-19-vaccine-demonstrates-893-efficacy-uk-phase-3).

At the beginning of the SARS-CoV-2 pandemic there was false optimism that the age structure of African populations may mitigate against severe continental impact. Our study does not uphold this conclusion. A wide range of now well-recognised co-morbidities (most commonly hypertension, type 2 diabetes mellitus and obesity) that associate with COVID-19 severity and death were present in the bulk of our cases and also in relevant other disease controls (ODC). Overall, 29% COVID-19 patients died. SARS-CoV-2 acutely brings into focus interaction between infectious and non-communicable conditions in Africa and the necessity that multimorbidity research must encompass such interaction much more in the future.

The analysis was not designed or powered to reproduce the previously documented relationships between HIV-1 co-infection and tuberculosis and risk of death from COVID-19 ^11^. The intent was rather to describe clinical presentation. Against an overall study mortality from COVID-19 of 28.8%, 19.4% of those HIV-1 co-infected, 40% of those with coincident tuberculosis, and 3/8 (37.5%) with coincident HIV-tuberculosis, died. Lymphocyte numbers were lowest, some inflammatory markers were higher and anti-SARS-CoV-2 antibody responses were most depressed in those exhibiting triple infection. The late detection of unsuspected tuberculosis in several otherwise typical COVID-19 patients is a cause for concern and increased clinical vigilance. We have documented here and elsewhere ^42^ that tuberculosis may adversely impact both antibody and T cell responses to SARS-CoV-2. Conversely, the immunological milieu set up in the lung by SARS-CoV-2, to which corticosteroid therapy will add, may have contributed to reactivation of subclinical or latent tuberculosis ^44^. Thus, in environments in which tuberculosis is common, clinical suspicion of reactivation and also that SARS-CoV-2 may co-exist in typical presentations of tuberculosis are the most important clinical lessons from our work.

## Data Availability

Data will be made available following formal publication via contact with the corresponding author

## Author contributions

RJW, EdB, CS, MC, AKC, KAW, AGD, RP-JL and CR conceived the study with input from JdV and NT on ethical aspects. MSM, GM, NN and SW supervised clinical recruitment. EdB, RD, CS recruited patients with logistic assistance from RTG. AJ designed the clinical research forms and database and supervised data cleaning. BWA conceptualised the percentage unaffected radiographic score. QSH performed the radiographic interpretation and scoring. FA co-ordinated the receipt and assay of laboratory specimens and MH was responsible for processing and reporting of samples in the NHLS analyses. EdB, CS and CR analyzed the data. All authors critically reviewed and provided input to the manuscript.

## Acknowledgments

We wish to thank all study participants and their relatives for their willingness to be recruited into the study. We thank the management of Groote Schuur Hospital for permitting the study to take place and its staff for their practical assistance in its conduct. We are grateful for logistic support of Berenice Arendse in the Institute of Infectious Disease and Molecular Medicine at the University of Cape Town.

## Funding

This research was funded in whole, or in part, by Wellcome [104803, 203135, 222754]. For the purpose of Open Access, the author has applied a CC BY public copyright licence to any Author Accepted Manuscript version arising from this submission. RJW was supported by the Francis Crick Institute which receives its core funding from Cancer Research UK (FC0010218), the UK Medical Research Council (FC0010218), and Wellcome (FC0010218).

## Copyright

The Corresponding Author has the right to grant on behalf of all authors and does grant on behalf of all authors, an exclusive licence (or non exclusive for government employees) on a worldwide basis to the BMJ Publishing Group Ltd to permit this article (if accepted) to be published in BMJ editions and any other BMJPGL products and sublicences such use and exploit all subsidiary rights, as set out in our licence.

## Competing interests

All authors have completed the <u>Unified Competing Interest form</u> (available on request from the corresponding author) and declare: no financial relationships with any organisations that might have an interest in the submitted work in the previous three years, no other relationships or activities that could appear to have influenced the submitted work

## Transparency declaration

The corresponding authors (the manuscript’s guarantors) affirm that the manuscript is an honest, accurate, and transparent account of the study being reported; that no important aspects of the study have been omitted; and that any discrepancies from the study as planned (and, if relevant, registered) have been explained.

## Notes

### Competing Interest Statement

The authors have declared no competing interest.

### Author Declarations

The study was approved by the University of Cape Town Health Sciences Research Ethical Committee (HREC 207/2020)

