## Supplement for "Communicable and non-communicable co-morbidities and the presentation of COVID-19 in an African setting of high HIV-1 and tuberculosis prevalence"

**Communicable and non-communicable co-morbidities and the presentation of SARS-CoV2 in an African setting of high HIV-1 and tuberculosis prevalence: supplemental data**

1. Complete list of HIATUS investigators.
2. Supplemental Table 1: Final diagnoses in non-COVID-19 control participants.
3. Supplemental Table 2: COVID-19 outcome in relation to presenting features.
4. Supplemental Table 3: Clinical features of COVID-19 participants with tuberculosis and HIV-tuberculosis.
5. Supplemental Table 4: Characteristics of HIV-1 uninfected and infected persons with either immune evidence of tuberculosis sensitization but no symptoms (latent tuberculosis, LTBI) or microbiologically-confirmed pulmonary tuberculosis recruited to previous studies.
6. Supplemental Figure 1: Type, number, and frequency and types of co-morbidity present in COVID-19 and ODC participants.
7. Supplementary Figure 2: Relationships between radiographic scores.
8. Supplemental Figure 3: Radiographic appearances of combined SARS-CoV2 and tuberculosis infection in the presence or absence of HIV-1 co-infection.
9. Supplemental Figure 4: Percentage of peripheral lymphocytes CD4 and CD8 positive in relation to COVID-19 severity and the presence or absence of HIV-1 and/or tuberculosis co-morbidities.
10. References.

**1 Complete list of HIATUS investigators**

Fatimah Abrahams, University of Cape Town

Brian Allwood, University of Stellenbosch

Saalikha Aziz, University of Cape Town

Nonzwakazi Bangani, University of Cape Town

John Black, Livingstone Hospital, Port Elizabeth

Melissa Blumenthal, University of Cape Town

Marise Bremer, University of Cape Town

Wendy Burgers, University of Cape Town

Maddalena Cerrone, University of Cape Town, Imperial College London and Francis Crick Institute

Zandile Ciko, University of Cape Town

Anna K Coussens, University of Cape Town and Walter and Eliza Hall Institute of Medical Research, and University of Melbourne

Remy Daroowala, Imperial College London and University of Cape Town

Angharad G Davis, Francis Crick Institute, University of Cape Town and University College London

Jantina de Vries, University of Cape Town

Elsa du Bruyn, University of Cape Town

Hanif G Esmail, University College London and University of Cape Town

Rene T Goliath, University of Cape Town

Siamon Gordon, University of Oxford

Yolande XR Harley, University of Cape Town

Marvin Hsiao, University of Cape Town

Amanda Jackson, University of Cape Town

Rachel P-J Lai, Imperial College London and Francis Crick Institute, London

Francisco Lakay, University of Cape Town

Fernando-Oneissi Martinez-Estrada, University of Surrey

Graeme Meintjes, University of Cape Town

Marc Mendelson, University of Cape Town

Ntobeko Ntusi, University of Cape Town

Tari Papavarnavas, University of Cape Town

Alize Proust, Francis Crick Institute, London

Catherine Riou, University of Cape Town

Sheena Ruzive, University of Cape Town

Qonita Said-Hartley, University of Cape Town

Georgia Schafer, International Centre for Genetic Engineering and Biotechnology, Cape Town

Keboile Serole, University of Cape Town

Cari Stek, Imperial College London and University of Cape Town

Nicki Tiffin, University of Cape Town

Sean Wasserman, University of Cape Town

Claire Whitaker, University of Cape Town

Katalin A Wilkinson, Francis Crick Institute and University of Cape Town

Robert J Wilkinson, University of Cape Town, Imperial College London and Francis Crick Institute

Kennedy Zvinairo, University of Cape Town

**Supplemental Table 1**

|  | **Disease** | **n** | **HIV-1**  **co-infected** |
| --- | --- | --- | --- |
| Non communicable disease (n=20) | Chronic cardiac failure | 7 | 0/7 |
|  | Exacerbation asthma / COPD | 6 | 1/6 |
|  | Diabetic keto-acidosis | 3 | 0/3 |
|  | Cerebrovascular accident | 1 | 0/1 |
|  | Gastro-intestinal | 2 | 1/2 |
|  | Pulmonary embolism | 1 | 0/1 |
|  | HIV-1 | 13 | - |
| Infectious diseases  (n=13) | Tuberculosis | 5 | 5/5 |
|  | *Pneumocystis jirovecii* pneumonia | 5 | 5/5 |
|  | Community acquired pneumonia | 2 | 0/2 |
|  | Urinary tract infection | 1 | 0/1 |
| Unclear final diagnosis at discharge^*^ |  | 2 | 0/2 |
| Outpatients from COVID testing centre (n=7) | Lower respiratory tract infection symptoms | 7 | 1/7 |

**Supplemental Table 1:** Final diagnosis of non-COVID-19 control participants.

^*^COVID-19 actively excluded

**Supplemental Table 2**

|  | **Survived**  **71.2%** (n=74) | **Died**  **28.8%** (n=30) | ***p-value*** |
| --- | --- | --- | --- |
| **Age** (median, IQR) | 52 [44-57] | 55 [46-66] | 0.10 |
| **Male** (%, n) | 54% (n=40) | 76.7% (n=23) | **0.032** |
| **HIV-1 co-infected** (%, n) | 33.8% (n=25) | 20% (n=6) | 0.16 |
| on antiretroviral therapy (ART) | 72% (n=18) | 83.3% (n=5) | 0.39 |
| Time on ART (years) ^a^ | 9.5 [6-12] | 10 [3.5-11] | 0.75 |
| CD4 count (cells/mm^3^) ^a^ | 144 [53-332] | 113 [45-270] | 0.71 |
| Log Viral load ^a^ | <1.3 [<1.3-4] | 3.17 [<1.3-5.21] | 0.42 |
| ***M. tuberculosis* positive** (%, n) | 12% (n=9) | 20% (n=6) | 0.30 |
| Previous tuberculosis episode/s (within 5 years) | 22.2% (n=2) | 33.3% (n=2) | >0.99 |
| **Co-morbidities** |  |  |  |
| Cardiovascular | 6.7% (n=5) | 6.7% (n=2) | 0.98 |
| Hypertension | 44.6% (n=33) | 56.7% (n=17) | 0.26 |
| Diabetes | 36.5% (n=27) | 46.7% (n=14) | 0.33 |
| Obesity | 29.3% (n=22) | 33.3% (n=10) | 0.36 |
| Other respiratory diseases | 9.2% (n=7) | - | - |
| **SARS-CoV-2 serology positive**^b^ | 67.6% (n=50) | 73.3% (n=22) | 0.56 |
| Cut-off index (median, IQR) | 5.5 [0.29-21.95] | 13.4 [0.43-35.6] | 0.094 |
| **WHO COVID-19 ordinal scale at enrolment (%, n)** |  |  |  |
| 3 | 22.6% (n=18) | - | - |
| 4 | 44% (n=33) | 17.2% (n=5) | **0.007** |
| 5 | 22.6% (n=17) | 44.8% (n=13) | **0.038** |
| 6 | 8.1% (n=6) | 36.7% (n=11) | **0.0004** |
| 7 | - | 3.4% (n=1) | - |
| **Severe (WHO** ≥**5)** | 31.1% (n=23) | 83.3% (n=25) | **<0.0001** |
| **Cycle threshold SARS PCR** (n=85) ^a^ | 31.5 [26.8-34] | 29.1 [25.4-33.6] | 0.34 |
| **On steroid treatment (%, n)** | 73% (n=54) | 93.3% (n=28) | **0.021** |
| **Overall days in clinical care** ^a^ | 11 [6-23] | 15 [7-22] | 0.54 |

**Supplemental Table 2:** COVID-19 Outcome in relation to presenting features.

IQR: interquartile range

^a^: Median and [IQR]

^b^: SARS-CoV-2 serology was performed using the Roche Elecsys assay, measuring SARS-CoV-2 nucleocapsid-specific antibodies. Results are reported as numeric values in form of a cut-off index (signal sample/cut-off), where a COI < 1.0 corresponds to non-reactive plasma and COI ≥ 1.0 to reactive plasma.

**Supplemental Table 3**

| **Age group** | **HIV-1 status** | **CD4 /mm^3^** | **Viral Load** | **on ART Y/N** | **Method of TB diagnosis** | **Time between + SARS CoV-2 PCR and TB dx** | **Prior TB episodes** | **Time since last TB episode** | **WHO score at enrolment** | **Outcome** |
| --- | --- | --- | --- | --- | --- | --- | --- | --- | --- | --- |
| 35-40 | -ve | NA | NA | NA | sputum Xpert + (Rif S) | simultaneous | 2 | 3 years | 4 | discharged |
| 40-45 | -ve | NA | NA | NA | sputum Xpert + (Rif S) | TB dx 25 days after | 0 | NA | 5 | discharged |
| 35-40 | -ve | NA | NA | NA | sputum culture + (after 24 days; sputum Xpert ND) | TB dx 6 weeks prior | 0 | NA | 3 | discharged |
| 40-45 | -ve | NA | NA | NA | sputum Xpert + (Rif S) | simultaneous | 0 | NA | 4 | discharged |
| 40-45 | -ve | NA | NA | NA | sputum Xpert + (Rif R) | simultaneous | 0 | NA | 6 | died |
| 60-65 | -ve | NA | NA | NA | sputum Xpert + (Rif S) | TB dx 20 days after | 0 | NA | 7 | died |
| 60-65 | -ve | NA | NA | NA | sputum Xpert + (Rif S) | simultaneous | 0 | NA | 6 | died |
| 30-35 | +ve | 106 | ND | N | pleural fluid Xpert + (Rif S) | simultaneous | 0 | NA | 3 | discharged |
| 30-35 | +ve | 110 | 17870 | Y | sputum Xpert + (Rif S) | TB dx 3 months prior | 0 | NA | 4 | discharged |
| 35-40 | +ve | 26 | 523463 | Y | clinical diagnosis: disseminated TB | TB dx 18 days prior | 1 | 2 years | 4 | died |
| 40-45 | +ve | 51 | 2941 | Y | sputum Xpert + (Rif S), sputum auramine 3+ | simultaneous | 1 | 4 months | 4 | died |
| 45-50 | +ve | 106 | 15860 | N | sputum Xpert + (Rif S) and culture positive (Rif S) | simultaneous | 0 | NA | 3 | discharged |
| 50-55 | +ve | 17 | 201574 | N | pericardial fluid and sputum Xpert sputum Xpert + (Rif S); urine LAM + | simultaneous | 0 | NA | 3 | discharged |
| 55-60 | +ve | 209 | LDL | Y | sputum Xpert + (Rif S) | TB dx 21 days after | 0 | NA | 5 | died |
| 25-30 | +ve | ND | 395 | Y | urine LAM positive, urine Mtb culture positive (TTP 20 days, Rif R, INH S) | simultaneous | 2 | 1 year | 3 | discharged |

**Supplemental Table 3:** Clinical features of COVID-19 participants with tuberculosis and HIV-tuberculosis.

Diagnoses of COVID-19 and TB occurring within 5 days of each other were denoted “simultaneous”.

+ve: positive, -ve: negative, dx: diagnosis, INH: isoniazid, LAM: Lipoarabinomannan, LDL: Lower than Detectable Limit, Mtb: *Mycobacterium tuberculosis*, NA: not applicable, ND: not done, Rif: Rifampicin, R: resistant, S: sensitive, TTP: time to positivity

**Supplemental Table 4**

|  | **HC (HIV-/aTB-)** | **HIV+/aTB-** | **HIV-/aTB+** | **HIV+/aTB+** |
| --- | --- | --- | --- | --- |
| n | 72 | 29 | 28 | 34 |
| Age ^a^ | 32 [26-38] | 34 [32-42] | 33 [28-47] | 37 [32-45] |
| Male (%) | 48.6% | 24.1% | 71.4% | 73.5% |
| CD4 count (cells/mm^3^) ^a^ | nd | 481 [358-700] | nd | 236 [121 - 355] |
| Log HIV viral load^a^ | na | <1.3 [<1.3-4.18] | na | 4.49 [2.19-5.00] |
| On ART (%) | na | 80.6 % | na | 38.9% |
| Unaffected Lung (%) ^a^ | na | na | 50 [30-70] | 60 [30-90] |
| CRP (µg mL^-1^) ^a^ | 1 [1-4] | 3 [2-10] | 100 [27.5-115] | 72 [36-123] |

**Supplemental Table 4:** Characteristics of healthy controls (HC) and mono- and co-infected HIV-1 (HIV+) and microbiologically confirmed pulmonary tuberculosis (TB+) controls, recruited to previous studies ^1 2^.

ART: Anti-retroviral treatment, CRP: C-reactive protein, nd: not done, na: not applicable.

^a^ Medians and [interquartile range]

**Supplemental Figure 1**

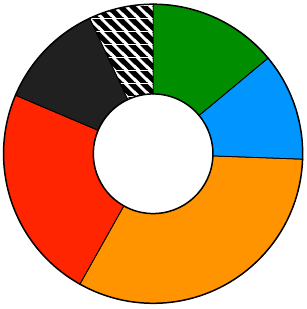

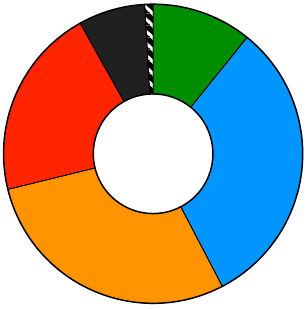

10.8%

31.5%

28.8%

20.7%

7.2

**n=104**

**n=42**

14%

11.6%

32.6%

23.2%

11.6%

7%

**Other disease controls**

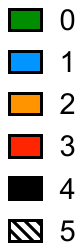

Hypertension (n=50/23)

Diabetes (n=41/12)

Obesity (n=34/14)

HIV-1 (n=31/13)

Active tuberculosis (n=15/5)

Cardiovascular (n=7/18)

Other respiratory (n=7/11)

Kidney disease (n=4/0)

Malignancy (n=3/0)

Immunosuppressive therapy (n=1/1)

Transplant recipient (n=1/0)

**Co-morbidities**

**COVID-19**

**Supplemental Figure 1**

Type, number, and frequency and types of co-morbidity present in COVID-19 and other disease control participants.

**Supplemental Figure 2**

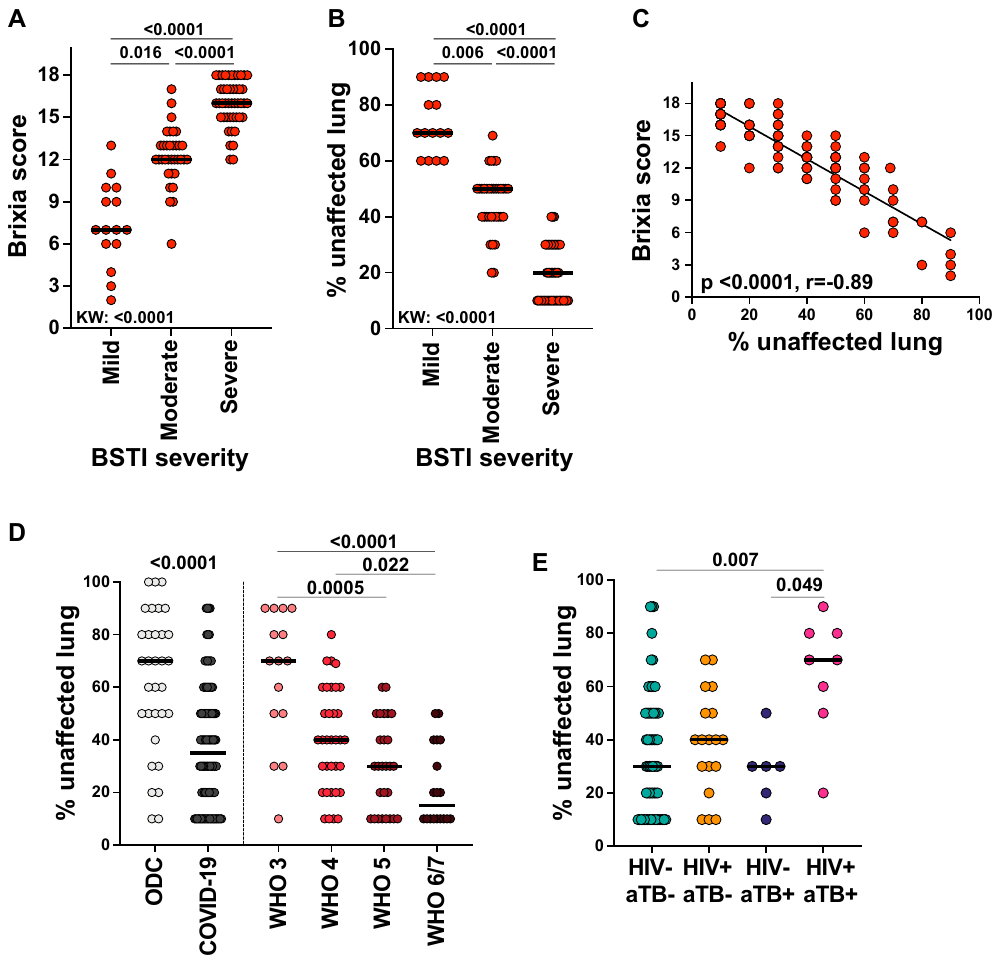

**Supplemental Figure 2 Relationships between radiographic scores**

**A** Relationship between Brixia and British Society for Thoracic Imaging (BSTI) radiographic severity scores ^3 4^.

**B** Inverse relationship between the BSTI radiographic severity score and the percentage

unaffected lung.

**C** Inverse correlation between percentage of lung unaffected and Brixia severity score. Non-parametric Spearman correlation.

**D** Percentage unaffected lung scores in non-COVID-19 hospitalized patients (ODC) and COVID-19 patients.

and relationship between percentage unaffected lung and WHO clinical severity score

**E** Relationship between the absence (-) or presence (+) of HIV-1 and/or tuberculosis (TB)

co-infection and percentage unaffected lung in COVID-19 patients.

Statistical comparisons were performed by the Kruskal-Wallis test (with Dunn's multiple comparison adjustments) in A, B, E and for the comparison of unaffected lung scores between patients grouped by WHO score. The comparison of OCD vs COVID-19 in D was done using a the Mann-Witney test.

**Supplemental Figure 3**

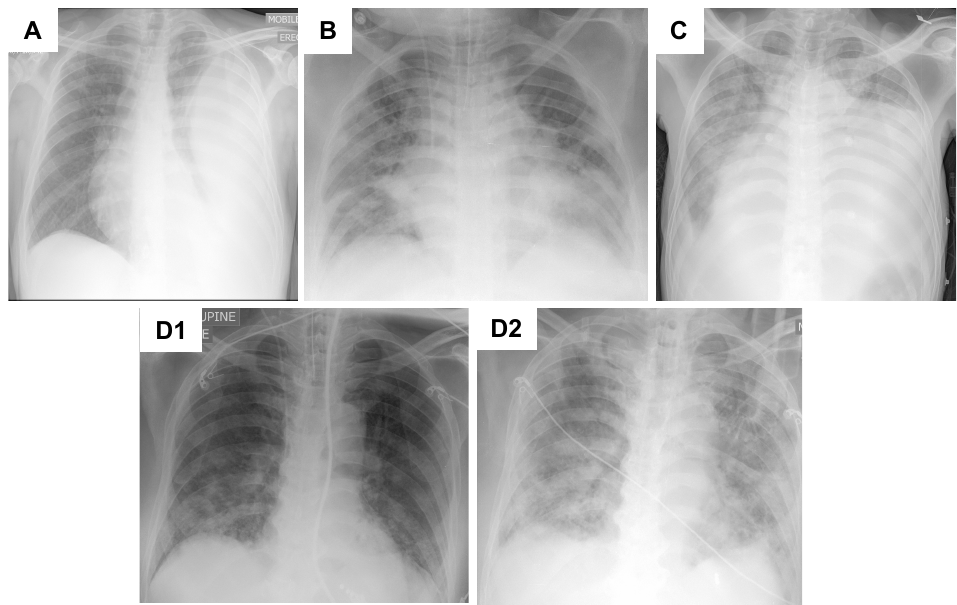

**Supplemental Figure 3 Radiographic appearances of combined SARS-CoV2 and tuberculosis infection in the presence or absence of HIV-1 co-infection**

A. 30-35-year-old HIV-1 infected antiretroviral-naive patient with a CD4 count of 106 cells/mm^3^ presenting with a large left pleural effusion that yielded a positive Gene Xpert MTB/Rif result. Also, SARS-CoV-2 RT-PCR positive with threshold cycle of 23.03. Uneventful course and discharged after 15 days on antitubercular therapy.
B. 40-45-year-old HIV-1 uninfected patient with hypertension, obesity and type II diabetes mellitus. SARS-CoV-2 RT-PCR positive and WHO grade 6. Admitted to intensive care where a tracheal aspirate was Gene Xpert TB/Rif positive with rifampin resistance detected. The patient had a complicated course with *Candida albicans* and *Serratia marcescens s*uperinfections and died after 21 days.
C. 50-55-year-old HIV-1 infected antiretroviral-naive patient with a CD4 count of 17 cells/mm^3^ presenting with a large pericardial effusion that was Gene Xpert MTB/Rif test positive. Also respiratory tract SARS-CoV-2 RT-PCR test was positive. Pericardiocentesis was performed and the patient was commenced on antitubercular therapy and discharged well to stepdown care after 7 days’ admission.
D. 60-65-year-old HIV-1 uninfected patient with hypertension, obesity and type II diabetes mellitus presented ketoacidotic and in severe respiratory failure. SARS-CoV-2 RT-PCR positive. Intubated and ventilated. By day 25 of intensive care admission, radiographic deterioration prompted a Gene Xpert MTB/Rif test which was positive. The patient developed multiorgan failure and died on day 28 of admission.

**Supplemental Figure 4**

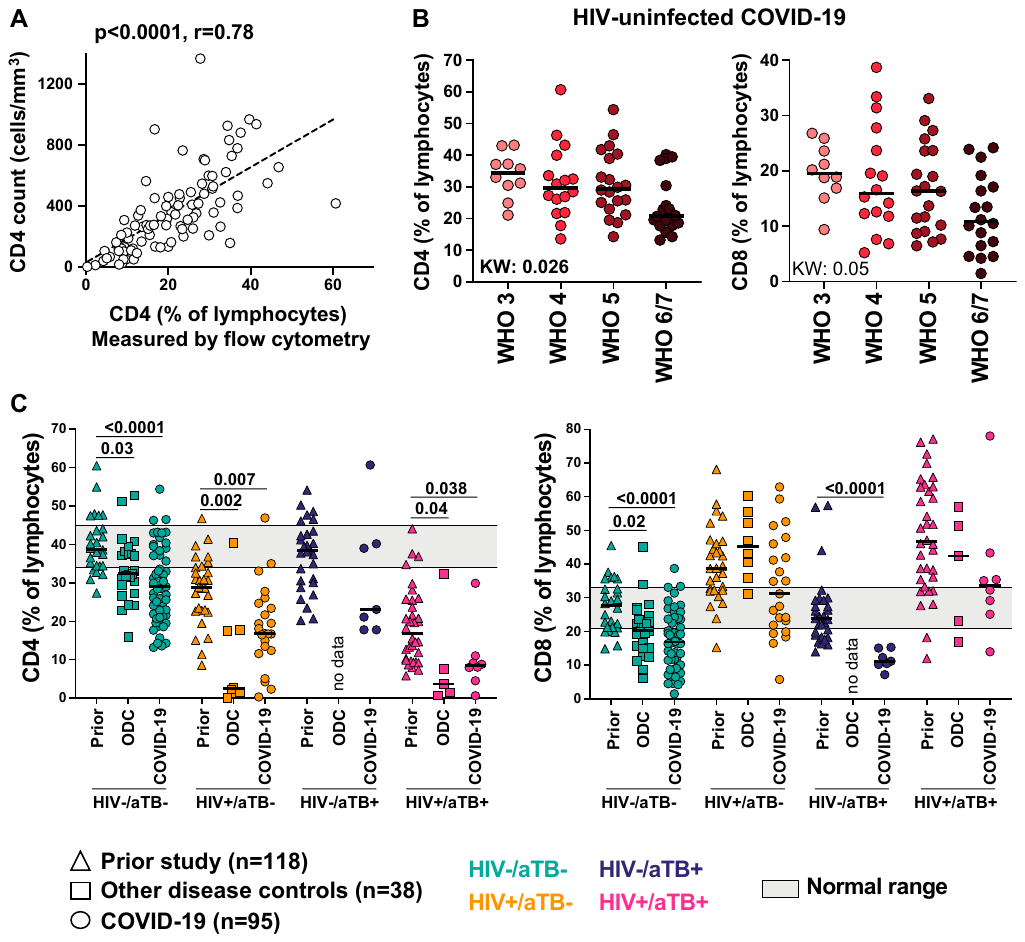

**Supplemental Figure 4 Percentage of peripheral lymphocytes CD4 and CD8 positive in relation to COVID-19 severity and the presence or absence of HIV-1 and/or tuberculosis co-morbidities**

**A.** Correlation between absolute CD4 lymphocyte count determined by coulter analysis and the CD4 percentage determined by flow cytometric analysis. Correlation was tested by a two-tailed non-parametric Spearman rank test.

**B**. Amongst HIV-1 uninfected patients with COVID-19 there was a trend towards a lower percentage of lymphocytes positive for CD4 (B) and CD8 T cells with increasing WHO grade severity, which was significant for CD4 (Kruskal Wallis test).

**C**. Additional control values were obtained from a subset of participants enrolled to a prior study of 118 ambulant HIV-1 uninfected and infected persons with either immune evidence of tuberculosis sensitization but no symptoms or microbiologically-confirmed pulmonary tuberculosis. When compared to HIV-1 uninfected healthy persons, the percentage of lymphocytes positive for CD4 was lower in both HIV-1 uninfected ODC and more so in COVID-19 patients. The pattern was reversed amongst HIV-1 infected patients with CD4 lymphocytes being especially low for ODC. Amongst patients with coincident HIV-1 and tuberculosis infection, % CD4 was very low irrespective of the presence or absence of SARS-CoV2 infection. When compared to HIV-1 uninfected healthy persons, the percentage of lymphocytes positive for CD8 was also lower in both HIV-1 uninfected ODC and more so in COVID-19 patients. However, this pattern was not observed amongst HIV-1 infected patients. The % CD8 positive lymphocyte was markedly depressed in HIV-1 uninfected patients with coincident tuberculosis and SARS-CoV2 infection.

Comparisons between groups were performed using a Kruskal-Wallis test.
